# Causal associations between fatty acid measures and schizophrenia – a two-sample Mendelian randomization study

**DOI:** 10.1101/2021.03.17.21253783

**Authors:** Hannah J Jones, Maria Carolina Borges, Rebecca Carnegie, David Mongan, Peter J Rogers, Sarah J Lewis, Andrew D Thompson, Stanley Zammit

## Abstract

**Objective:** Although studies suggest that erythrocyte concentrations of omega-3 and omega-6 fatty acids are lower in individuals with schizophrenia, evidence of beneficial effects of omega-3 fatty acid supplementation is limited. This study therefore aimed to determine whether omega-3 and omega-6 fatty acid levels are causally related to schizophrenia.

**Methods:** Causality was evaluated using the inverse variance weighted (IVW) 2-sample Mendelian randomization (MR) method using fatty acid levels and schizophrenia genome-wide association study results. Weighted median, weighted mode, and MR Egger regression methods were used as sensitivity analyses. To address the mechanism, analyses were performed using instruments within the FADS and ELOVL2 genes. Multivariable MR (MVMR) was used to estimate direct effects of omega-3 fatty acids on schizophrenia, independent of omega-6 fatty acids, lipoproteins and triglycerides.

**Results:** MR analyses indicated that long-chain omega-3 and omega-6 fatty acid levels were associated with lower risk of schizophrenia (docosahexaenoic acid [DHA] ORIVW: 0.83, 95% CI: 0.75-0.92). In contrast, short-chain fatty acids were associated with an increased risk of schizophrenia (alpha-linolenic acid ORIVW: 1.07, 95% CI: 0.98-1.18). Causal effects were consistent across sensitivity and FADS single-SNP analyses. MVMR indicated that the protective effect of DHA on schizophrenia persisted after conditioning on other lipids (ORIVW: 0.84, 95% CI: 0.71-1.01).

**Conclusions:** Results are consistent with protective effects of long-chain omega-3 and omega-6 fatty acids on schizophrenia suggesting that people with schizophrenia may have difficulty converting short-chain to long-chain PUFAs. Long-chain PUFA supplementation or diet enrichment, particularly in higher risk individuals, might help prevent onset of disorder.

## Introduction

Schizophrenia is a serious psychiatric disorder that affects approximately 0.7% of the population (1) and places a substantial burden on sufferers and the healthcare system (2). Development of novel prevention or treatment interventions has been limited by the paucity of robust evidence of potentially modifiable risk factors.

Polyunsaturated fatty acids (PUFAs) are key components of cellular and intracellular membranes implicated in normal brain development and function (3), and are mainly divided into omega-3 fatty acids and omega-6 fatty acids. These are derived from alpha-linolenic acid (ALA, 18:3n-3) and linoleic acid (LA, 18:2n-6), respectively, through a succession of elongation, desaturation, and β-oxidation events involving competition for shared enzymes and metabolic substrates. Omega-3 fatty acids, especially docosahexaenoic acid (DHA, 22:6n-3), play vital roles in maintaining cell membrane integrity and fluidity which in turn have effects on processes including the functioning of membrane proteins and cellular signalling (4).

Decreased concentrations of PUFAs have been characterised in erythrocyte membranes in individuals with schizophrenia (5, 6) and at ultra-high risk for psychosis (7, 8). Findings are primarily based on relatively small case-control studies that are subject to selection bias, reverse causation and confounding, and meta-analyses report considerable heterogeneity of results (5, 6). Even studies showing altered erythrocyte membrane PUFA levels in ultra-high risk or first-episode psychosis individuals when not on antipsychotic medication (7, 8) cannot rule out reverse causation as a result of poor diet secondary to illness or confounding by social or other dietary factors.

Randomised-controlled trials (RCTs) of omega-3 fatty acid supplements in individuals with schizophrenia have provided little evidence of benefits in psychotic symptom reduction (9-12). Furthermore, whilst a beneficial and long-term effect of omega-3 supplementation reducing transition to psychosis was reported in an RCT of 81 individuals at ultra-high risk for psychosis (13, 14), a subsequent, larger RCT showed limited evidence to support this (15). Arguments have been put forward as to why the effect size for the main outcome in this latter trial may have been under-estimated, and in a post-hoc analysis, increases in erythrocyte omega-3 fatty acids were associated with reduced symptom severity and better functioning (16). However, it is difficult to be confident that i) PUFAs have causal effects on psychosis, and ii) if so, which PUFA might be most important to supplement to negate this effect.

RCTs are expensive and time-consuming, therefore alternative methods to strengthen causal inference from observational data could help guide whether such trials are potentially warranted. Mendelian randomization (MR) is a causal inference method which, if certain assumptions are met, can overcome issues of reverse causation and confounding that weaken inferences from observational analyses by using genetic variants as instrumental variables (17) (see Supplement). We therefore aimed to apply a 2-sample MR approach to determine whether omega-3 and omega-6 fatty acid levels are causally related to schizophrenia.

## Methods

This study utilised deidentified summary-level, publicly available data (see Table S1 in the online supplement for data sources and sample sizes). Ethical approval was obtained in all original studies.

### Fatty acid genetic instruments

Results from a meta-analysis of 10 fatty acid genome-wide association studies (GWASs; units = standard deviation [sd] mmol/L) (18) were used to generate instruments for circulating levels of DHA and LA. Single nucleotide polymorphism (SNPs) that did not reach genome-wide significant evidence of association (p ≤ 5e-8) or had a minor allele frequency ≤ 0.01 were excluded.

Following harmonisation of the fatty acid SNP instrument information with the corresponding outcome data (see below) and linkage disequilibrium (LD) clumping using the TwosampleMR R package (v0.5.5) (19), instrument sets contained 5 independent SNPs for DHA and 12 independent SNPs for LA (Supplementary Methods, Table S2 in the online supplement), which were then used to instrument omega-3 and omega-6 fatty acid levels, respectfully (see below).

In a complementary and mechanistically informative analysis, instruments were identified for the FADS gene cluster and ELOVL2 gene which encode key fatty acid desaturase and elongase enzymes, respectively. SNPs within the FADS gene cluster locus (chr11:61560452-61659523 ± 500kb) and ELOVL2 gene locus (chr6:10980992-11044624 ± 500kb) were extracted from the DHA and LA fatty acid GWAS summary statistics and LD clumped, which left 1 SNP to represent the FADS gene cluster (rs174546) and 1 SNP to represent the ELOVL2 gene (rs2281591).

### Fatty acid exposure data

Due to sample overlap between the fatty acid GWAS used to identify our instruments and the outcome schizophrenia GWAS, fatty acid effect sizes for use in the MR analyses were extracted from fatty acid GWASs with no sample overlap with the schizophrenia GWAS.

Information for the 5 omega-3, 12 omega-6, FADS gene cluster and ELOVL2 gene instruments was extracted from results of the Cohorts for Heart and Aging Research in Genomic Epidemiology (CHARGE) Consortium GWAS of plasma levels of omega-3 fatty acids (ALA, eicosapentaenoic acid [EPA; 20:5n-3], docosapentaenoic acid [DPA, 22:5n-3], DHA) (20) and omega-6 fatty acids (LA, arachidonic acid [AA; 20:4n-6], adrenic acid [AdA, 22:4n-6]) (21). Before analysis, CHARGE effect estimates (% of total fatty acids) were converted to standardised units (sd mmol/L) using GWIS (“Genome-wide Inferred Study”) (22) (Supplementary Methods).

### Schizophrenia outcome data

For each fatty acid instrument SNP, the corresponding SNP information (namely SNP alleles, effect allele frequency, effect size [log odds ratio], standard error and p-value) was extracted from the most recent schizophrenia GWAS (23).

### Two-sample Mendelian randomization

#### Univariable Mendelian randomization analyses

For each fatty acid instrument set, SNP-fatty acid and SNP-schizophrenia data were harmonised and 2-sample MR analyses performed using the TwosampleMR R package (v0.5.5). Within our primary analyses, the inverse variance weighted (IVW) method (17) was used to estimate causal effects. The weighted median (24), weighted mode (25), and MR Egger (26) methods were used in sensitivity analyses as each make different assumptions regarding instrument validity. Details are available in the Supplementary Methods.

For the FADS gene cluster and ELOVL2 gene single SNP analyses, ratio estimates were calculated by dividing the SNP-schizophrenia effect estimate by the SNP-fatty acid effect estimate with standard errors derived using the first term from a delta method expansion for the ratio estimate (27).

#### Multivariable Mendelian randomization analyses

##### Omega-3 and omega-6 fatty acids

Multivariable MR (MVMR) (28) was performed to simultaneously estimate the direct effects of omega-3 and omega-6 fatty acid levels on schizophrenia. MVMR analyses focused on fatty acids at equivalent positions on the biosynthesis pathway, namely estimating ALA effects while conditioning on LA, and EPA effects while conditioning on AA.

Prior to MVMR analyses, the 5 omega-3 and 12 omega-6 instruments used in the IVW analyses were combined and further LD clumped, to ensure that SNPs were independent. This resulted in a set of 14 MVMR instruments. SNP information for these 14 SNPs was extracted from the CHARGE omega-3 and omega-6 fatty acid GWAS summary statistics (20, 21) and harmonised with the outcome schizophrenia GWAS information as previously described. Multivariable IVW effect estimates were then derived using the MVMR R package (28).

##### Omega-3 fatty acids and lipid measures

As the majority of fatty acid instruments fall into loci that are also associated with lipoprotein and triglyceride phenotypes (Table S2 in the online supplement), MVMR was used to estimate the direct causal effect of DHA and LA levels independent of effects of high-density lipoprotein (HDL), low-density lipoprotein (LDL) and triglycerides (TGs) on schizophrenia.

Genetic associations between SNPs and LDL, HDL, and TGs were obtained from a recent GWAS carried out in a UK Biobank sample of 403,943-441,016 individuals of European descent (29) which identified 534, 220 and 440 genome-wide significant loci associated with HDL, LDL and/or TGs, respectively. These SNPs were merged with the instruments for DHA and LA separately, LD clumped and harmonised with the schizophrenia outcome data. The final instrument sets contained 112 SNPs for the DHA MVMR analysis and 116 SNPs for the LA MVMR analysis. SNP information for these instrument sets was extracted from the CHARGE omega-3 and omega-6 fatty acid, UK Biobank Lipid, and schizophrenia GWAS summary statistics (20, 21, 29) and multivariable IVW effect estimates were derived.

### Assessing instrument strength and effect heterogeneity

Instrument strength was quantified using the mean F statistic 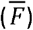 (30) within the univariable IVW analyses, the conditional F statistic (28, 31) within the multivariable IVW analyses, and the total variance explained by each multi-SNP instrument set (Supplementary Methods).

The degree of violation of the IVW and MR-Egger assumption that the SNP-exposure association is measured without error (the NO Measurement Error’ [NOME] assumption) was assessed using 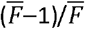 for IVW analyses and the I2GX statistic for MR Egger analyses (30), with values close to 1 indicating minimal regression dilution bias due to violation of the NOME assumption (30, 32).

Presence of heterogeneity between individual SNP-outcome/SNP-exposure effect estimates was assessed using Cochrane’s (IVW) and Rücker’s (MR Egger) Q tests (32, 33).

## Results

### Instrument strength and effect heterogeneity

Within the univariable analyses, 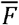 statistics ranged from 19.12-175.47 indicating that IVW estimates were not likely subject to weak instrument bias, as was the case for the ALA and LA MVMR analysis (Table 3). However, the conditional F-statistics indicated probable weak instrument bias for both the EPA and AA MVMR and the fatty acid and lipid levels MVMR analyses.

The total variance explained (R2) by each multi-SNP instrument set used within the multi-locus MR analyses was between 1.08-9.27% for the DHA instruments and omega-3 fatty acid measures and between 6.47-16.50% for the LA instruments and omega-6 fatty acid measures (Table S3 in the online supplement). With the exception of the univariable DHA analyses in which the I2GX statistic was low (I2GX = 0.02), statistics referring to the degree of violation of the NOME assumption indicated that measurement error in the SNP-exposure associations was not substantially attenuating the effect estimates 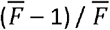 ranged from 0.95-0.99 and I2GX ranged from 0.85-0.99) (Table S3 in the online supplement).

There was some evidence of heterogeneity between each SNP causal effect estimates in the univariable ALA, EPA, DPA, and AA analyses (Table S3 in the online supplement).

### Univariable Mendelian randomization

Multi-locus MR analyses indicated that long-chain omega-3 and omega-6 fatty acid levels, that is fatty acids derived following elongation and desaturation of ALA and LA precursors, were associated with lower risk of schizophrenia (e.g. DHA: IVW odds ratio [OR]: 0.83, 95% confidence interval [CI]: 0.75-0.92) (Table 1). In contrast, ALA and LA were associated with an increased risk of schizophrenia (e.g. ALA: IVW OR: 1.07, 95% CI: 0.98-1.18) (Table 1). Estimated causal effects were consistent across the other MR methods that make different assumptions about pleiotropy (Table 1).

**Table 1.**
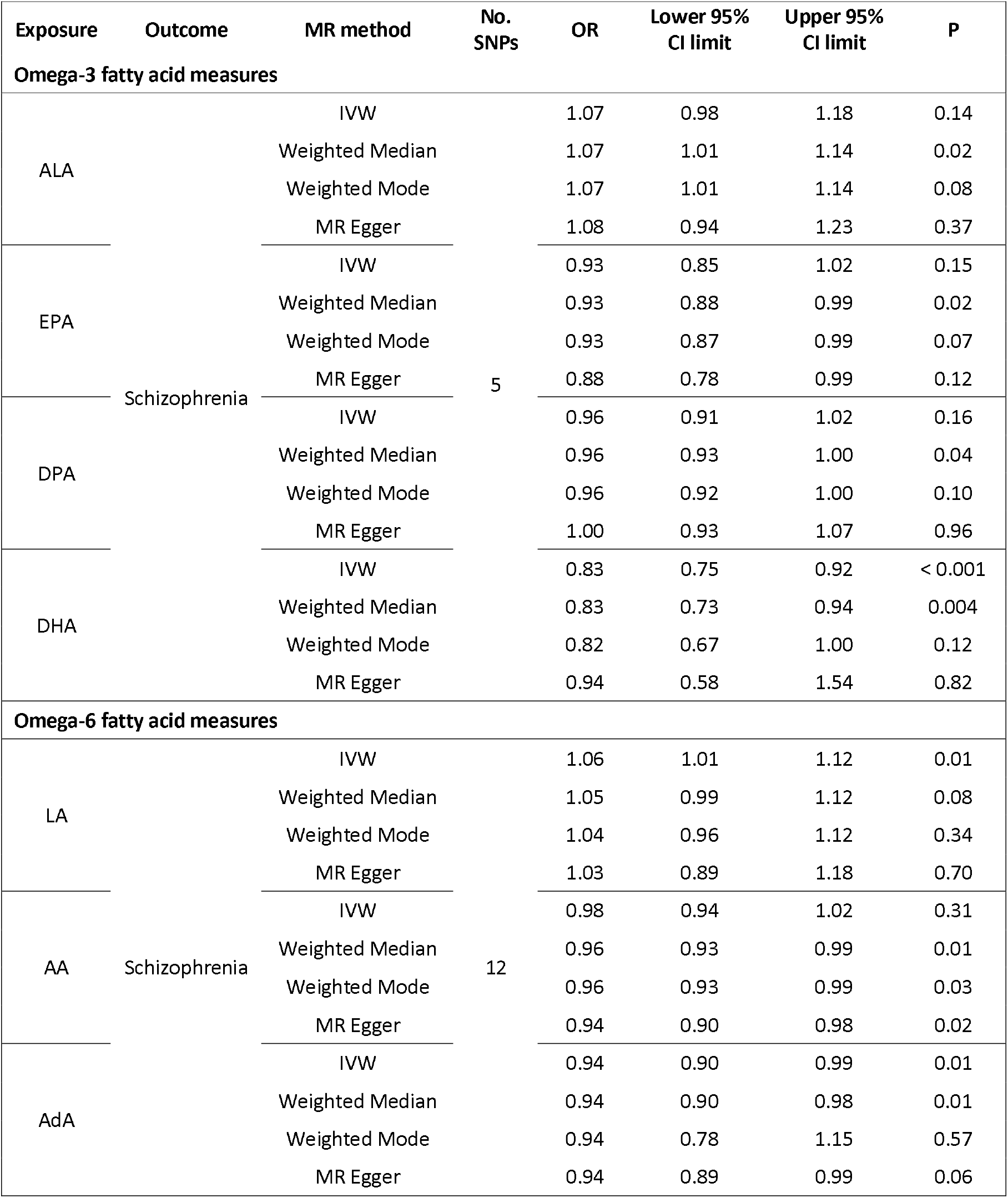
Univariable causal effects of fatty acids on schizophrenia. Odds ratios of schizophrenia are presented per standard deviation (mmol/L) increase in serum fatty acid measure as estimated by univariable Mendelian randomization methods. MR, Mendelian randomization; No. SNPs, number of single nucleotide polymorphism used in the analysis as instruments; OR, odds ratio; 95% CI, 95% confidence interval; IVW, inverse variance weighted; ALA, alpha-linolenic acid; EPA, eicosapentaenoic acid; DPA, docosapentaenoic acid; DHA, docosahexaenoic acid; LA, linoleic acid; AA, arachidonic acid; AdA, adrenic acid

Except for the analysis of AA, the MR Egger intercept test provided little evidence for the presence of pleiotropic effects across the multi-locus analyses (Table S4 in the online supplement). There was also slight asymmetry in the funnel plots of instrument precision for the EPA, DPA, DHA, and AA analyses which is indicative of directional pleiotropy (Figures S2-S4,S6 in the online supplement).

With the exception of analyses of DHA and LA, leave-one-out analyses indicated that IVW results were being driven by SNPs within the FADS gene cluster (Figures S1-7). Estimates from the FADS single-SNP analyses also indicated that long-chain omega-3 and omega-6 fatty acid levels were associated with lower risk of schizophrenia (EPA OR: 0.92, 95% CI: 0.86-0.98; DPA OR: 0.95, 95% CI: 0.91-0.99; DHA OR: 0.74, 95% CI: 0.58-0.96; AA OR: 0.96, 95% CI: 0.93-0.99; AdA OR: 0.95, 95% CI: 0.91-0.99) and the short-chain omega-3 and omega-6 fatty acid levels were associated with an increased risk of schizophrenia (ALA OR: 1.08, 95% CI: 1.02-1.15; LA OR: 1.18, 95% CI: 1.04-1.36) (Table 2).

**Table 2.** Causal effects of fatty acids on schizophrenia via the FADS gene cluster and ELOVL2. Odds ratios of schizophrenia are presented per standard deviation (mmol/L) increase in serum fatty acid measures estimated using a single SNP instruments within the FADS gene cluster and ELOVL2. No. SNPs, number of single nucleotide polymorphism used in the analysis as instruments; OR, odds ratio; 95% CI, 95% confidence interval; ALA, alpha-linolenic acid; EPA, eicosapentaenoic acid; DPA, docosapentaenoic acid; DHA, docosahexaenoic acid; LA, linoleic acid; AA, arachidonic acid; AdA, adrenic acid.

| Exposure | Outcome | OR | Lower 95%<br>CI limit | Upper 95% CI<br>limit | P |
| --- | --- | --- | --- | --- | --- |
| *FADS* gene cluster (rs174546) |  |  |  |  |  |
| Omega-3 fatty acid measures |  |  |  |  |  |
| ALA | Schizophrenia | 1.08 | 1.02 | 1.15 | 0.01 |
| EPA |  | 0.92 | 0.86 | 0.98 | 0.01 |
| DPA |  | 0.95 | 0.91 | 0.99 | 0.01 |
| DHA |  | 0.74 | 0.58 | 0.96 | 0.02 |
| Omega-6 fatty acid measures |  |  |  |  |  |
| LA | Schizophrenia | 1.18 | 1.04 | 1.36 | 0.01 |
| AA |  | 0.96 | 0.93 | 0.99 | 0.01 |
| AdA |  | 0.95 | 0.91 | 0.99 | 0.01 |
| *ELOVL2* (rs2281591) |  |  |  |  |  |
| Omega-3 fatty acid measures |  |  |  |  |  |
| ALA | Schizophrenia | 1.48 | 0.16 | 13.48 | 0.73 |
| EPA |  | 1.04 | 0.88 | 1.21 | 0.66 |
| DPA |  | 1.02 | 0.94 | 1.1 | 0.66 |
| DHA |  | 0.96 | 0.78 | 1.17 | 0.66 |
| Omega-6 fatty acid measures |  |  |  |  |  |
| LA | Schizophrenia | 1.18 | 0.55 | 2.51 | 0.68 |
| AA |  | 1.09 | 0.74 | 1.59 | 0.66 |
| AdA |  | 1.06 | 0.81 | 1.39 | 0.66 |

**a** Estimated by dividing the SNP-schizophrenia effect estimate by the SNP-fatty acid effect estimate with standard errors derived using the first term from a delta method expansion for the ratio estimate (27)

In contrast, estimates from the ELOVL2 single-SNP analyses were more imprecise and compatible with both risk-increasing and protective effects for each of the fatty acid measures (Table 2).

### Multivariable Mendelian randomization

When the associations between schizophrenia and ALA and LA levels were assessed together using IVW MVMR, effect estimates for ALA were comparable to the univariable IVW estimates (ALA univariable OR: 1.07, 95% CI: 0.98-1.18; ALA multivariable OR: 1.08, 95% CI: 0.99-1.17). However, evidence of a risk increasing effect of LA on schizophrenia weakened (LA univariable OR: 1.06, 95% CI: 1.01 −1.12; LA multivariable OR: 0.98, 95% CI: 0.92-1.04) (Table 3).

Effect estimates from MVMR analyses of EPA and AA were consistent with the univariable analyses, though were less precise (Table 3).

**Table 3.** Multivariable Mendelian randomization analyses estimating the direct effects of omega-3 fatty acids conditioning on omega-6 fatty acids. IVW OR, inverse variance weighted odds ratio; 95% CI, 95% confidence interval; ALA, alpha-linolenic acid; LA, linoleic acid; EPA, eicosapentaenoic acid; AA, arachidonic acid.

| Exposure | Outcome | No. SNP Instruments | No. SNPs in analysis | Conditional *F* statistic | IVW OR | Lower 95% CI limit | Upper 95% CI limit | P |
| --- | --- | --- | --- | --- | --- | --- | --- | --- |
| ALA | Schizophrenia | 5 | 14<sup>a</sup> | 20.63 | 1.08 | 0.99 | 1.17 | 0.12 |
| LA |  | 12 |  | 25.22 | 0.98 | 0.92 | 1.04 | 0.52 |
| EPA |  | 5 | 14<sup>a</sup> | 6.43 | 0.97 | 0.84 | 1.12 | 0.68 |
| AA |  | 12 |  | 6.89 | 0.97 | 0.91 | 1.04 | 0.45 |

<sup>a</sup> Less than the total number of instruments as there is overlap between the instrument sets.

IVW MVMR analyses estimating the direct effect of DHA levels on schizophrenia conditioned on HDL, LDL and TG levels, were comparable to the univariable IVW estimate (DHA univariable OR: 0.83, 95% CI: 0.75-0.92; DHA multivariable OR: 0.84, 95% CI: 0.71-1.01) and showed a protective effect of DHA on schizophrenia (Table 4). IVW MVMR analyses estimating the direct effect of LA levels conditioned on HDL, LDL and TG levels no longer provided evidence of a risk increasing effect of LA on schizophrenia (LA univariable OR: 1.06, 95% CI: 1.01 −1.12; LA multivariable OR: 0.96, 95% CI: 0.91-1.06) (Table 4).

**Table 4.** Multivariable Mendelian randomization analyses estimating the direct effects of omega-3 and omega-6 fatty acids conditioning on lipid measures. IVW OR, inverse variance weighted odds ratio; 95% CI, 95% confidence interval; DHA, docosahexaenoic acid; HDL, high density lipoprotein; LDL, low density lipoprotein; TG, triglycerides; LA, linoleic acid.

| Exposure | Outcome | No. SNP Instruments | No. SNPs in analysis | Conditional *F* statistic | IVW OR | Lower 95% CI limit | Upper 95% CI limit | P |
| --- | --- | --- | --- | --- | --- | --- | --- | --- |
| DHA | Schizophrenia | 5 | 112<sup>a</sup> | 1.53 | 0.84 | 0.71 | 1.01 | 0.07 |
| HDL |  | 75 |  | 6.83 | 1.00 | 0.87 | 1.15 | 0.96 |
| LDL |  | 35 |  | 2.66 | 1.02 | 0.85 | 1.23 | 0.83 |
| TG |  | 56 |  | 4.66 | 0.92 | 0.77 | 1.09 | 0.31 |
| LA | Schizophrenia | 12 | 116<sup>a</sup> | 2.62 | 0.96 | 0.91 | 1.06 | 0.39 |
| HDL |  | 74 |  | 6.97 | 0.98 | 0.89 | 1.08 | 0.64 |
| LDL |  | 45 |  | 3.33 | 0.97 | 0.88 | 1.08 | 0.61 |
| TG |  | 56 |  | 4.32 | 1.04 | 0.91 | 1.19 | 0.57 |

<sup>a</sup> Less than the total number of instruments as there is overlap between the instrument sets.

## Discussion

In this study we aimed to use MR to strengthen the causal inferences that can be drawn about the effect of omega-3 and omega-6 fatty acids on schizophrenia risk. We observed that schizophrenia risk was associated with higher levels of the short-chain, more saturated PUFAs, ALA and LA. In contrast, we observed that higher levels of the long-chain, less saturated PUFAs, EPA, DPA, DHA, AA and AdA, were associated with a lower risk of schizophrenia, with the strongest evidence being for a protective effect of the omega-3 fatty acid DHA. Our leave-one-out and single-locus analyses indicate that these effects were primarily driven by SNPs in the FADS gene cluster, and hence desaturation steps during PUFA biosynthesis might play a key role in this relationship. The exception to this was for DHA, where results do not appear to be driven solely through variants within the FADS gene cluster. We also showed that the effect of DHA on schizophrenia is similar when conditioning on other lipids, indicating that the effects of PUFA are likely to be independent of (and not mediated or confounded by) HDL, LDL or triglycerides.

MR can provide evidence on causal effects of exposures if key assumptions relating to instrument validity are met. Our results were consistent across different MR methods that make different assumptions about horizontal pleiotropy, suggesting that horizontal pleiotropy is unlikely to be an adequate explanation for our findings. There was also little evidence of pleiotropy from the MR Egger intercept estimates (except for AA), although these estimates are likely to be underpowered given the small number of instruments used in the current study. This also made subjective interpretation of asymmetry in funnel plots of instrument precision difficult and so rigorous assessment of the presence of directional, horizontal pleiotropy was not always possible.

The direct effect estimates of PUFA on schizophrenia were less precise (and evidence of causal effects weaker) in the MVMR analyses, most likely due to a decrease in power in these conditional analyses, especially for the long-chain PUFAs AA and EPA where instruments were weaker. Nevertheless, it appears that the omega-3 fatty acid biosynthetic pathway may be more important than the omega-6 one in relation to schizophrenia risk as the conditioned estimate for ALA was relatively unchanged, whilst that for LA was substantially attenuated.

Overall, the consistency of effect sizes across different methods, the strength of evidence, and our secondary analyses indicate that our findings are consistent with a causal effect of PUFA biosynthesis on schizophrenia. It is difficult to tease out, in the absence of trials, whether causal effects on schizophrenia result primarily from higher levels of short-chain PUFA or lower levels of long-chain ones as differences at any point along the PUFA biosynthesis pathway will lead to changes in both. For example, reduced function of the desaturase or elongase enzymes that regulate the introduction of double bonds and extend the fatty acyl chain, respectively, will result in accumulation of upstream PUFA and paucity of downstream ones. Furthermore, as the synthesis of both omega-3 and omega-6 fatty acids involve shared enzymes, it is difficult to disentangle independent effects from confounding, i.e,. a change in function of the elongase enzyme would lead to a change in levels of both long-chain omega-3 and omega-6 fatty acids, even if the causal effect is only due to a change in one of these.

Although our study provides evidence that higher levels of long-chain PUFA reduce the risk of developing schizophrenia, most RCTs of PUFA supplementation show little evidence of improved symptomatology in schizophrenia (9-12). Furthermore, the recent NEURAPRO trial in individuals at ultra-high risk of psychosis found no effect of supplements in reducing transition to psychosis (15).

Evidence using MR of causal effects of PUFA on schizophrenia needs to be interpretated as the risk from lifetime exposure to omega-3 and omega-6 fatty acids, or at least long-term exposure that might include particularly important periods of exposure in early development. Indeed, causal effects identified using MR could even be the result of maternal genetic effects, which are partly shared with the offspring, influencing intrauterine exposure to fatty acids during foetal development. As such, even if causal, PUFA supplements given over a relatively short-term in adulthood, as per recent RCTs, may not necessarily have any measurable impact. More specific MR frameworks, which aim to account for shared maternal and offspring genetic variants, could help to determine the importance of intrauterine exposure to PUFA on schizophrenia risk (34).

MR studies can also help to inform which supplements might be most likely to impact on risk. RCTs of PUFA supplementation in schizophrenia or ultra-high risk samples have used a variety of different PUFA and at different ratios, with most using EPA (or its ester) or a combination of EPA and DHA. As can be seen from our study, protective effects of PUFA increase as PUFA chain lengthens, with strongest evidence present for the omega-3 fatty acid, DHA. As genetic instruments for PUFA and schizophrenia continue to improve, MR studies could shed further light on the most important PUFA to supplement in trials.

Finally, although our results indicate that omega-3 fatty acids may be more likely to be causally related to schizophrenia than omega-6 fatty acids, our results are consistent with causal effects of both. If independent causal effects of both omega-3 and omega-6 are present, this might argue against a primarily inflammation-based explanation for PUFA effect on psychosis given that these seem to have opposite effects on inflammation (35), with omega-3 fatty acids tending to have anti-inflammatory and omega-6 pro-inflammatory properties (36). Whilst the methods of action of PUFA on schizophrenia are likely to be more complex than this, teasing out independent effects could inform mechanistic pathways in schizophrenia aetiology.

### Strengths and limitations

The strengths of this study are that we use an instrumental variable analysis that is unlikely to be affected by reverse causation and confounding when compared to traditional observational designs. Further, we use the largest GWAS samples available to identify our instruments and model the outcome. We also employ different methodological approaches to estimate causal effects that allow us to be more confident that results are not due to horizontal pleiotropy.

However, our results must be interpreted in the context of a number of limitations. First, although we use the largest schizophrenia GWAS sample available, thus minimising measurement error bias in our outcome, due to issues of sample overlap we used instrument-exposure effect estimates from smaller, less powered fatty acid GWASs. Though the F-statistics for our univariable analyses did not indicate that our instruments were weak, there may be weak instrument bias in some of our MVMR analyses which limited our ability to tease out the direct effects of EPA while conditioning on AA.

Second, we were constrained in our ability to test a more comprehensive range of PUFA measures as we were only able to obtain GWAS data on the measures included. For example, although we used MVMR analyses to estimate the direct effects of one fatty acid while accounting for another, we were unable to estimate the causal effect of the omega-6:omega-3 fatty acid ratio that may be important in schizophrenia pathogenesis (37).

Third, the PUFA GWAS effect sizes were based on plasma levels and thus might not accurately reflect the membrane levels that might more strongly influence risk of schizophrenia through their involvement in synaptic membrane modification, brain cell signalling, modification of receptor properties and signal transduction (38, 39).

Finally, given the difficulties inherent in determining causal effects of diet on psychopathology, as evidenced by the disparate views regarding effects of PUFA on depression (40), a higher level of evidence is likely required to shift the argument regarding PUFA effects on psychosis.

### Conclusion

In this study, we have used MR to investigate whether PUFA have a causal effect on schizophrenia. Our results are consistent with protective effects of long-chain omega-3, and to a lesser extent, omega-6 fatty acids on schizophrenia. These findings suggest that people with schizophrenia are more likely to have difficulty converting short-chain to long-chain PUFAs, and that provision of the latter through supplementation or diet enrichment, particularly in higher risk individuals, might help prevent onset of disorder.

## Data Availability

This study utilised deidentified summary-level, publicly available data.

http://www.computationalmedicine.fi/data#NMR_GWAS.

http://www.chargeconsortium.com/main/results

http://www.chargeconsortium.com/main/results

https://www.med.unc.edu/pgc/download-results/

## Acknowledgments

This study was supported by the NIHR Biomedical Research Centre at University Hospitals Bristol and Weston NHS Foundation Trust and the University of Bristol. The views expressed are those of the authors and not necessarily those of the NIHR or the Department of Health and Social Care. DM is a Fellow on the Irish Clinical Academic Training (ICAT) Programme which is supported by the Wellcome Trust and the Health Research Board (Grant Number 203930/B/16/Z), the Health Service Executive National Doctors Training and Planning and the Health and Social Care, Research and Development Division, Northern Ireland. MCB is supported by a UK MRC Skills Development Fellowship (Grant Number MR/P014054/1). RC is supported by the Wellcome Trust (Grant Number WT 212557/Z/18/Z).

