## Supplementary material for "Causal associations between fatty acid measures and schizophrenia – a two-sample Mendelian randomization study"

### Supplementary Materials

#### Methods

##### Generating fatty acid genetic instruments

Single nucleotide polymorphism (SNP) instrument sets were generated for each fatty acid phenotype of interest, namely circulating levels of docosahexaenoic acid (DHA, 22:6n-3) and linoleic acid (LA, 18:2n-6), using results from a meta-analysis of 10 fatty acid genome-wide association studies (GWASs; up to 13,544 individuals of European descent, units = standard deviation [sd] mmol/L) (1). SNPs were included in each instrument set if they had a minor allele frequency (MAF) ≥ 0.01 and met genome-wide significant evidence of association ($p \leq 5e^{-8}$) within the fatty acid GWASs. Full GWAS summary statistics are available from: http://www.computationalmedicine.fi/data#NMR_GWAS.

Using the TwosampleMR R package (v0.5.5) (2), fatty acid SNP instrument information (SNP alleles, effect allele frequency) was harmonised with the outcome schizophrenia GWAS information to ensure SNP-exposure and SNP-outcome effects corresponded to the same allele. During harmonisation, palindromic SNPs with allele frequencies > 0.42 and < 0.58 or SNPs with mismatching alleles (not due to strand differences) were removed. To identify independent SNPs, harmonised SNP lists were clumped for linkage disequilibrium (LD) using the 2sampleMR R package "clump_data" function (2), *p*-values from each fatty acid measure GWAS, $r^2$ of 0.001, and a 10,000kb clumping window. This resulted in instrument sets containing 5 independent SNPs for DHA and 12 independent SNPs for LA that were also present within the outcome data (Table S1).

In a complementary and mechanistically informative analysis, an instrument was also identified for the *FADS* gene cluster and *ELOVL2* gene which encode key desaturase and elongase enzymes, respectively, involved in omega-3 and omega-6 fatty acid biosynthesis pathways. SNPs within the *FADS* gene cluster locus (chr11:61560452-61659523 ± 500kb) and *ELOVL2* gene locus (chr6:10980992-11044624 ± 500kb) were extracted from the DHA and LA fatty acid GWAS summary statistics (1). The *FADS* gene cluster and *ELOVL2* gene SNPs were LD clumped (using DHA GWAS *p*-values, and the parameters as described above) which left 1 genome-wide significant SNP to represent the *FADS* gene cluster (rs174546) and 1 genome-wide significant SNP to represent the *ELOVL2* gene (rs2281591).

#### Converting CHARGE GWAS effect estimates

Cohorts for Heart and Aging Research in Genomic Epidemiology (CHARGE) Consortium original effect estimates (expressed as % of total fatty acids) were converted to standard units of mmol/L (sd mmol/L) using GWIS ("Genome-wide Inferred Study") (3), a method that provides an approximation of GWAS summary statistics for a variable that is a function of phenotypes for which GWAS summary statistics, phenotypic means, and covariances are available. SNP-trait effect estimates were derived as a linear function of the allele frequencies, population means of the fatty acid concentrations (assumed to approximate the intercepts of the model) and SNP-fatty acid effect estimates. Corresponding standard errors were derived using the Delta-method having obtained the covariance matrix for effect estimates.

#### Multi-locus, 2-sample Mendelian randomization methods

Mendelian randomization (MR) is a causal inference method which, if certain assumptions are met, can overcome issues of reverse causation and confounding that weaken inferences from observational analyses by using genetic variants as instrumental variables (4, 5). The core assumptions of MR are i) the genetic instrumental variables must be associated with the risk factor of interest, ii) they share no common cause with the outcome (i.e., are independent of confounders), and iii) they only affect the outcome through the risk factor.

Two-sample MR is an extension of MR that allows the instrument-exposure and instrument-outcome associations to be measured in two independent samples (6), which is advantageous in the absence of large samples with measures of the exposure and outcome of interest, and can be implemented using summary data from large scale genome-wide association studies (GWASs) (7), providing an opportunity to substantially increase statistical power.

For each fatty acid instrument set, SNP-fatty acid and SNP-schizophrenia data were harmonised and 2-sample MR analyses performed using the TwosampleMR R package (v0.5.5) (2). Within our primary analyses, the inverse variance weighted (IVW) method was used to estimate causal effects. In addition, the weighted median, weighted mode, and MR Egger methods were used in sensitivity analyses as each make different assumptions regarding MR instrument validity.

Briefly, the IVW method is equivalent to a weighted linear regression of SNP-outcome associations on SNP-exposure associations with a constrained zero intercept. The IVW provides a robust causal estimate assuming that all SNPs are valid instruments, i.e., there is no directional, horizontal pleiotropy (where an IV affects the outcome through a different biological pathway from the exposure under investigation), or if present, the directional, horizontal pleiotropy is balanced (5, 7). The weighted median method estimates the causal effect from the median of the weighted empirical density function of individual SNP effect estimates (8) and provides a robust estimate even if up to 50% of the information in the analysis is from invalid SNPs. The weighted mode method estimates the causal effect from the mode of the weighted empirical density function of SNP effect estimates and provides a robust estimate assuming that the weights associated with valid instruments are the largest among all subsets of instruments (the ZEro Modal Pleiotropy Assumption) (9). The MR Egger regression method is an expansion of the IVW method that does not assume that all instruments are valid. Like the IVW, this method is a weighted linear regression, however the intercept is not constrained to zero (10). The method therefore provides a causal estimate that takes pleiotropic effects into account with the intercept giving an estimate of the average pleiotropic effect. The MR Egger method gives a valid causal estimate if the SNP-exposure associations are not correlated to the direct effects of the genetic variants on the outcome (i.e. pleiotropic effects) (the Instrument Strength Independent of Direct Effect (InSIDE) assumption) (10).

#### Assessing instrument strength

Instrument strength was quantified using the mean F statistic ($\overline{F}$) (11) within the univariable IVW analyses and the conditional *F* statistic (12, 13) within the multivariable IVW analyses. As knowledge of the covariance between the effect of the SNP on each exposure is needed to estimate the MVMR conditional *F* statistic when using exposure data from overlapping samples (12), covariance matrices were provided using the "phenocov_mvmr" function of the MVMR R package. For the lipid MVMR analyses, covariance matrices were generated using the phenotypic correlations reported in the lipid GWAS (14). For the total omega-3 and total omega-6 fatty acid MVMR analysis and sensitivity analyses, phenotypic correlations were estimated using the GWAS summary data and the PhenoSpD R toolkit (15).

The total variance explained ($R^2$) by each multi-SNP instrument set used within the regression-based MR analyses was calculated using the standardised GWAS effect size ($\beta$) and MAF for each SNP $j$ using the following equation (16):

$$R^2 = \sum_j 2{\beta_j}^2 \times MAF_j \times (1 - MAF_j)$$

#### Supplementary Tables

**Table S1. Data sources**

| Use in current study | Phenotype | Reference | N | Ancestry | Link to data source |
|---|---|---|---|---|---|
| Identifying instruments for fatty acid levels | Serum/plasma levels of DHA and LA | Kettunen et al. (1) | up to 13,544 | European | http://www.computationalmedicine.fi/data#NMR_GWAS. |
| Instrument-exposure information | Plasma levels of ALA, EPA, DPA and DHA | Lemaitre et al. (17) | up to 8,866 | European | http://www.chargeconsortium.com/main/results |
| Instrument-exposure information | Plasma levels of LA, AdA and AA | Guan et al. (18) | up to 8,631 | European | http://www.chargeconsortium.com/main/results |
| Instrument-outcome information | Schizophrenia | Schizophrenia Working Group of the PGC (19) | 69,369 cases, 236,642 controls | European (80%), East Asian (20%) | https://www.med.unc.edu/pgc/download-results/ |

**Note:** ALA, alpha-linolenic acid; EPA, eicosapentaenoic acid; DPA, docosapentaenoic acid; DHA, docosahexaenoic acid; LA, linoleic acid; AA, arachidonic acid; AdA, adrenic acid; PGC, Psychiatric Genomics Consortium

**Table S2. Fatty acid Instruments used for univariable, multi-locus Mendelian randomization analyses**

| Fatty acid measure | SNP | CHR | BP | Gene(s) † |
|---|---|---|---|---|
| DHA | rs2281591 | 6 | 10990493 | *ELOVL2* |
| | rs174546 | 11 | 61569830 | *FADS1, FADS2* |
| | rs11604424 | 11 | 116651115 | *ZPR1* |
| | rs261334 | 15 | 58726744 | *LIPC, ALDH1A2, LIPC-AS1* |
| | rs16996148 | 19 | 19658472 | *CILP2* |
| LA | rs11591147 | 1 | 55505647 | *PCSK9* |
| | rs7534572 | 1 | 62999675 | *DOCK7, AL451044.1* |
| | rs1260326 | 2 | 27730940 | *GCKR* |
| | rs4296389 | 2 | 21142994 | - |
| | rs79225634 | 5 | 74619639 | *AC008897.2* |
| | rs964184 | 11 | 116648917 | *ZPR1, AP006216.1* |
| | rs99780 | 11 | 61596633 | *FADS1, FADS2* |
| | rs174418 | 15 | 58687603 | *ALDH1A2* |
| | rs1800588 | 15 | 58723675 | *LIPC, ALDH1A2, LIPC-AS1* |
| | rs17231506 | 16 | 56994528 | *CETP* |
| | rs7412 | 19 | 45412079 | *APOE, AC011481.3* |
| | rs76366838 | 19 | 45399896 | *TOMM40* |

**Note:** SNP, single nucleotide polymorphism; CHR, chromosome; BP, base-pair
† Mapped genes determined by the Ensembl Variant Effect Predictor (20)

**Table S3. Statistics used to assess instrument strength and causal effect heterogeneity**

| Exposure | Outcome | Cochran's Q (P-value) | Rücker's Q (P-value) | $\overline{F}$ | $(\overline{F} - 1) / \overline{F}$ | $I^2_{GX}$ | *Variance explained* |
|---|---|---|---|---|---|---|---|
| **Omega-3 fatty acid measures** | | | | | | | |
| ALA | Schizophrenia | 9.86 (0.04) | 9.85 (0.02) | 63.10 | 0.98 | 0.98 | 3.44% |
| EPA | | 10.04 (0.04) | 5.91 (0.12) | 60.42 | 0.98 | 0.98 | 3.32% |
| DPA | | 10.32 (0.04) | 5.58 (0.13) | 175.47 | 0.99 | 0.99 | 9.27% |
| DHA | | 3.05 (0.55) | 2.81 (0.42) | 19.12 | 0.95 | 0.02 | 1.08% |
| **Omega-6 fatty acid measures** | | | | | | | |
| LA | Schizophrenia | 15.18 (0.17) | 14.78 (0.14) | 46.89 | 0.98 | 0.85 | 6.47% |
| AA | | 21.47 (0.03) | 12.36 (0.26) | 131.66 | 0.99 | 0.99 | 16.50% |
| AdA | | 14.94 (0.19) | 14.93 (0.13) | 53.75 | 0.98 | 0.98 | 6.98% |

**Note:** ALA, alpha-linolenic acid; EPA, eicosapentaenoic acid; DPA, docosapentaenoic acid; DHA, docosahexaenoic acid; LA, linoleic acid; AA, arachidonic acid; AdA, adrenic acid

**Table S4. Evidence of average pleiotropic effect as estimated by MR Egger regression**

| Fatty acid measure | No. SNPs | MR Egger Intercept OR (95% CI) | P |
|---|---|---|---|
| **Omega-3 fatty acid measures** | | | |
| ALA | 5 | 1.00 (0.98, 1.02) | 0.97 |
| EPA | | 1.01 (1.00, 1.03) | 0.24 |
| DPA | | 0.99 (0.97, 1.00) | 0.21 |
| DHA | | 0.99 (0.95, 1.03) | 0.66 |
| **Omega-6 fatty acid measures** | | | |
| LA | 12 | 1.00 (0.99, 1.02) | 0.62 |
| AA | | 1.01 (1.00, 1.02) | 0.02 |
| AdA | | 1.00 (0.99, 1.01) | 0.93 |

**Note:** MR, Mendelian randomization; No. SNPs, number of single nucleotide polymorphism used in the analysis as instruments; OR, odds ratio; 95% CI, 95% confidence interval; ALA, alpha-linolenic acid; EPA, eicosapentaenoic acid; DPA, docosapentaenoic acid; DHA, docosahexaenoic acid; LA, linoleic acid; AA, arachidonic acid; AdA, adrenic acid

#### Supplementary Figures

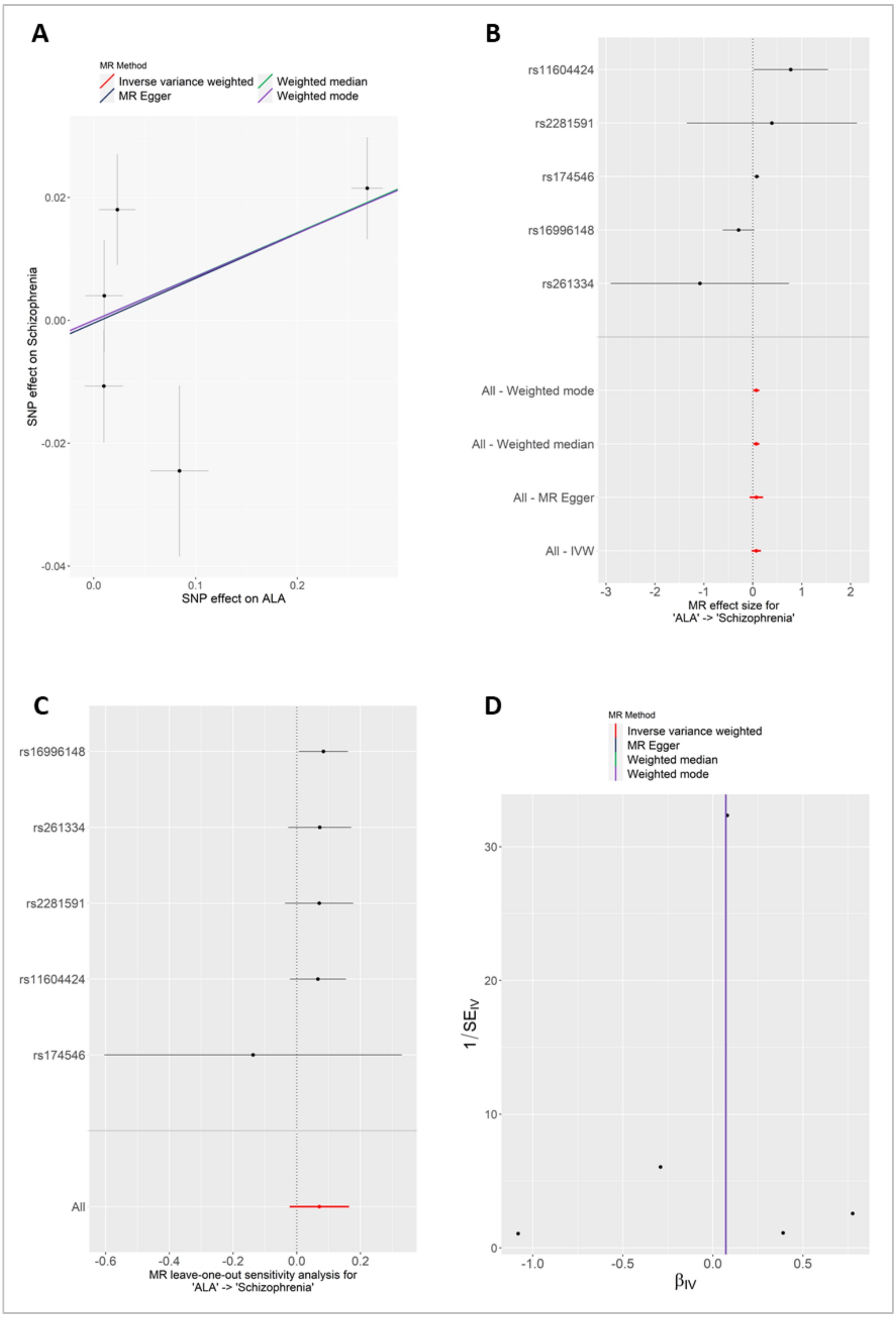


**Figure S1.** Mendelian randomization (MR) sensitivity plots of the causal effect of alpha-linolenic acid (ALA) levels on schizophrenia. A) Scatter plot showing results of the 4 MR multi-locus methods (see legend), B) Forest plot showing individual single nucleotide polymorphism (SNP) ratio estimates (SNP-outcome effect estimate / SNP-exposure effect estimate), C) Leave one out plot showing inverse variance weighted (IVW) estimates after omitting each SNP, and D) Funnel plot showing instrument precision: ratio estimate in log(odds ratio) ($\beta_{IV}$; x-axis) by instrument strength (1/ $\beta_{IV}$ standard error [SE]). Asymmetry in this plot may be a result of directional pleiotropy.

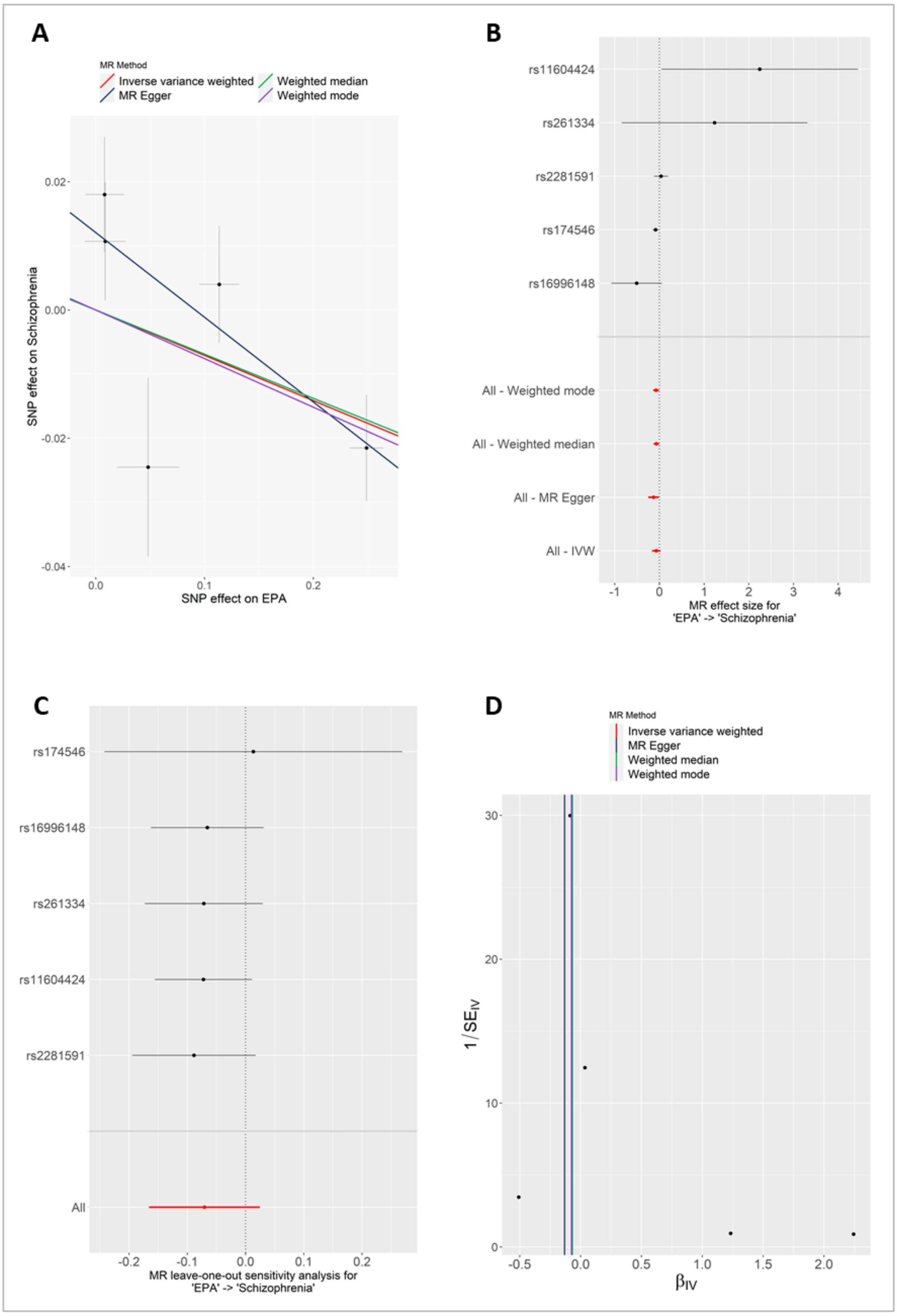


**Figure S2.** Mendelian randomization (MR) sensitivity plots of the causal effect of eicosapentaenoic acid (EPA) levels on schizophrenia. A) Scatter plot showing results of the 4 MR multi-locus methods (see legend), B) Forest plot showing individual single nucleotide polymorphism (SNP) ratio estimates (SNP-outcome effect estimate / SNP-exposure effect estimate), C) Leave one out plot showing inverse variance weighted (IVW) estimates after omitting each SNP, and D) Funnel plot showing instrument precision: ratio estimate in log(odds ratio) ($\beta_{IV}$; x-axis) by instrument strength (1/ $\beta_{IV}$ standard error [SE]). Asymmetry in this plot may be a result of directional pleiotropy.

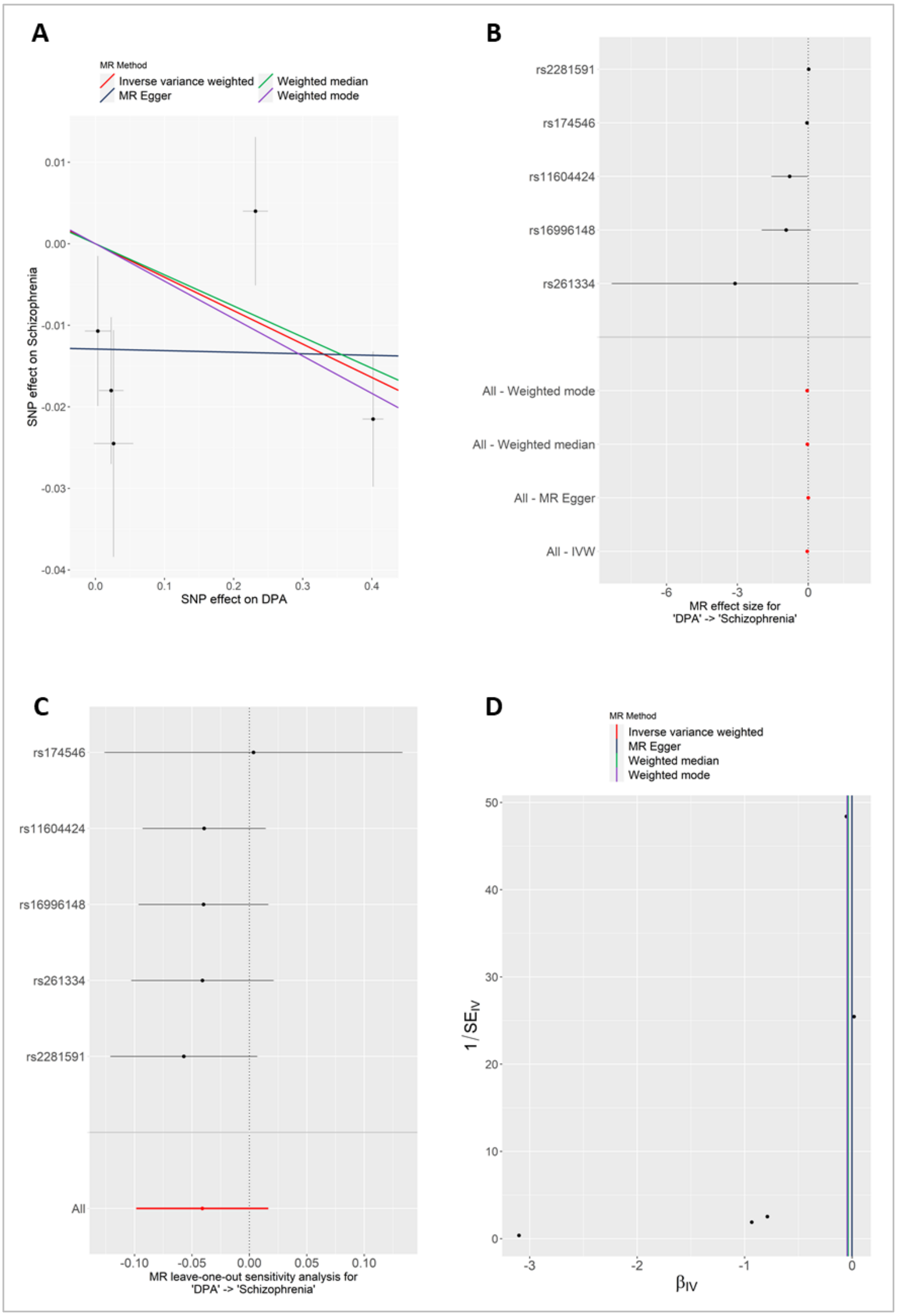


**Figure S3.** Mendelian randomization (MR) sensitivity plots of the causal effect of docosapentaenoic acid (DPA) levels on schizophrenia. A) Scatter plot showing results of the 4 MR multi-locus methods (see legend), B) Forest plot showing individual single nucleotide polymorphism (SNP) ratio estimates (SNP-outcome effect estimate / SNP-exposure effect estimate), C) Leave one out plot showing inverse variance weighted (IVW) estimates after omitting each SNP, and D) Funnel plot showing instrument precision: ratio estimate in log(odds ratio) ($\beta_{IV}$; x-axis) by instrument strength (1/ $\beta_{IV}$ standard error [SE]). Asymmetry in this plot may be a result of directional pleiotropy.

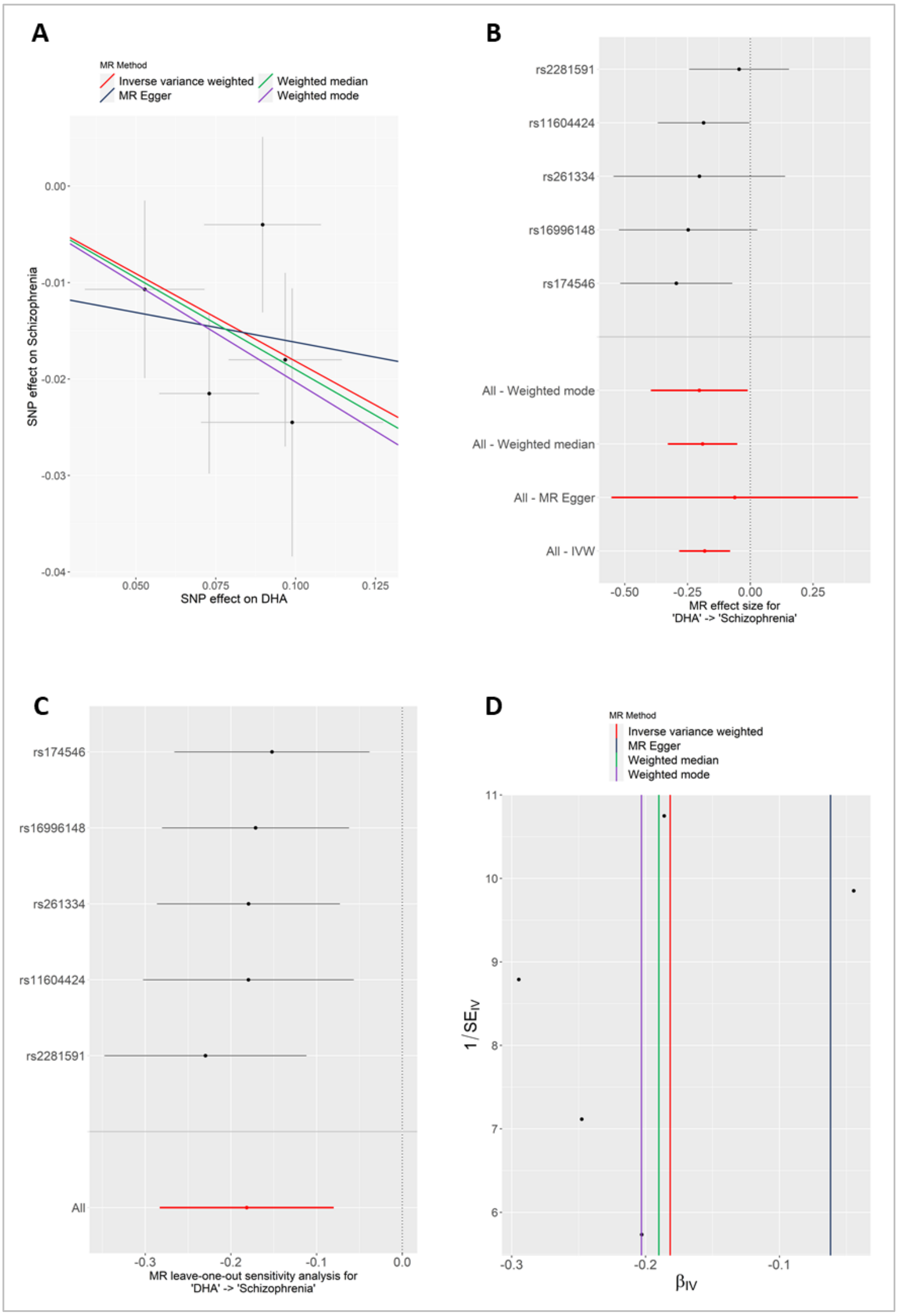


**Figure S4.** Mendelian randomization (MR) sensitivity plots of the causal effect of docosahexaenoic acid (DHA) levels on schizophrenia. A) Scatter plot showing results of the 4 MR multi-locus methods (see legend), B) Forest plot showing individual single nucleotide polymorphism (SNP) ratio estimates (SNP-outcome effect estimate / SNP-exposure effect estimate), C) Leave one out plot showing inverse variance weighted (IVW) estimates after omitting each SNP, and D) Funnel plot showing instrument precision: ratio estimate in log(odds ratio) ($\beta_{IV}$; x-axis) by instrument strength (1/ $\beta_{IV}$ standard error [SE]). Asymmetry in this plot may be a result of directional pleiotropy.

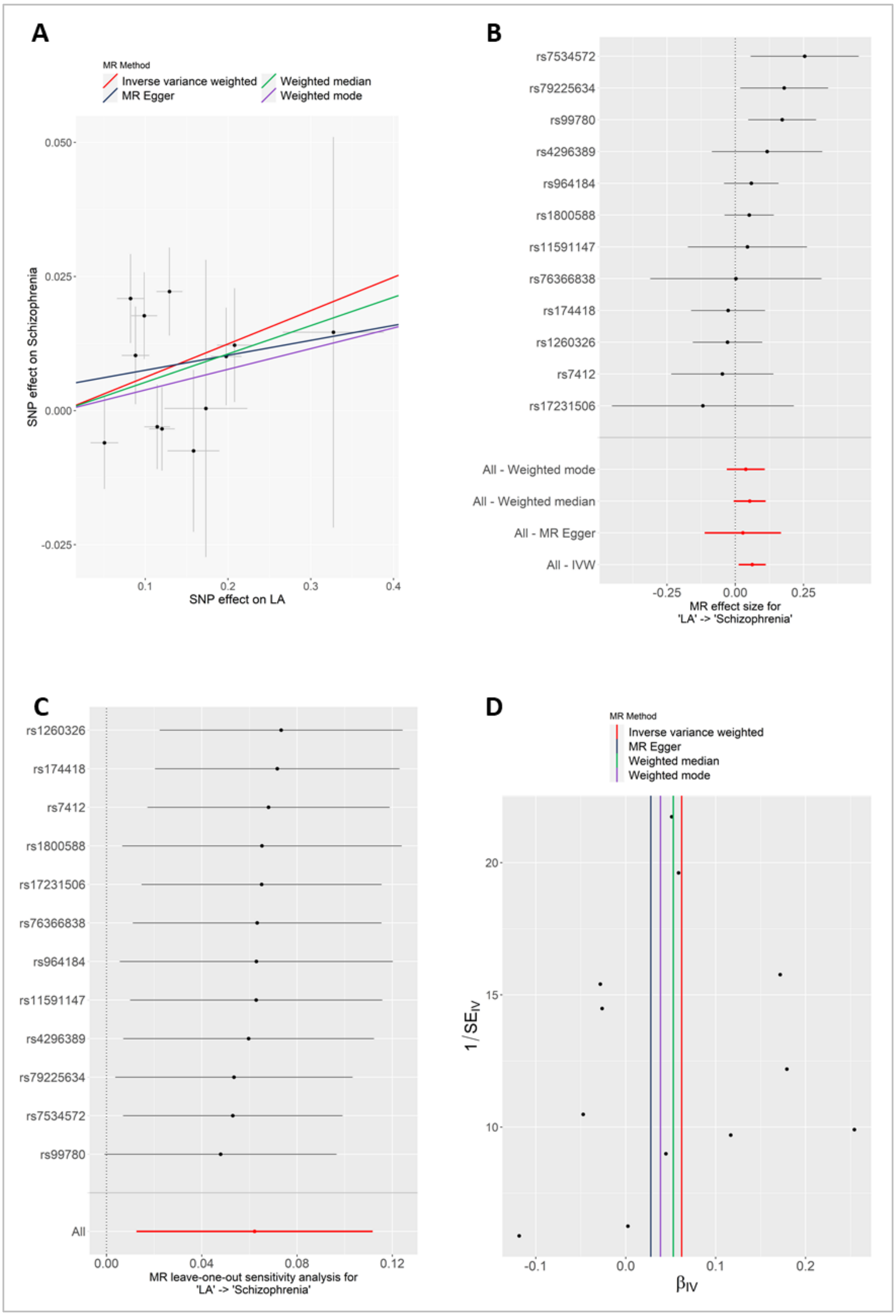


**Figure S5.** Mendelian randomization (MR) sensitivity plots of the causal effect of linoleic acid (LA) levels on schizophrenia. A) Scatter plot showing results of the 4 MR multi-locus methods (see legend), B) Forest plot showing individual single nucleotide polymorphism (SNP) ratio estimates (SNP-outcome effect estimate / SNP-exposure effect estimate), C) Leave one out plot showing inverse variance weighted (IVW) estimates after omitting each SNP, and D) Funnel plot showing instrument precision: ratio estimate in log(odds ratio) ($\beta_{IV}$; x-axis) by instrument strength (1/ $\beta_{IV}$ standard error [SE]). Asymmetry in this plot may be a result of directional pleiotropy.

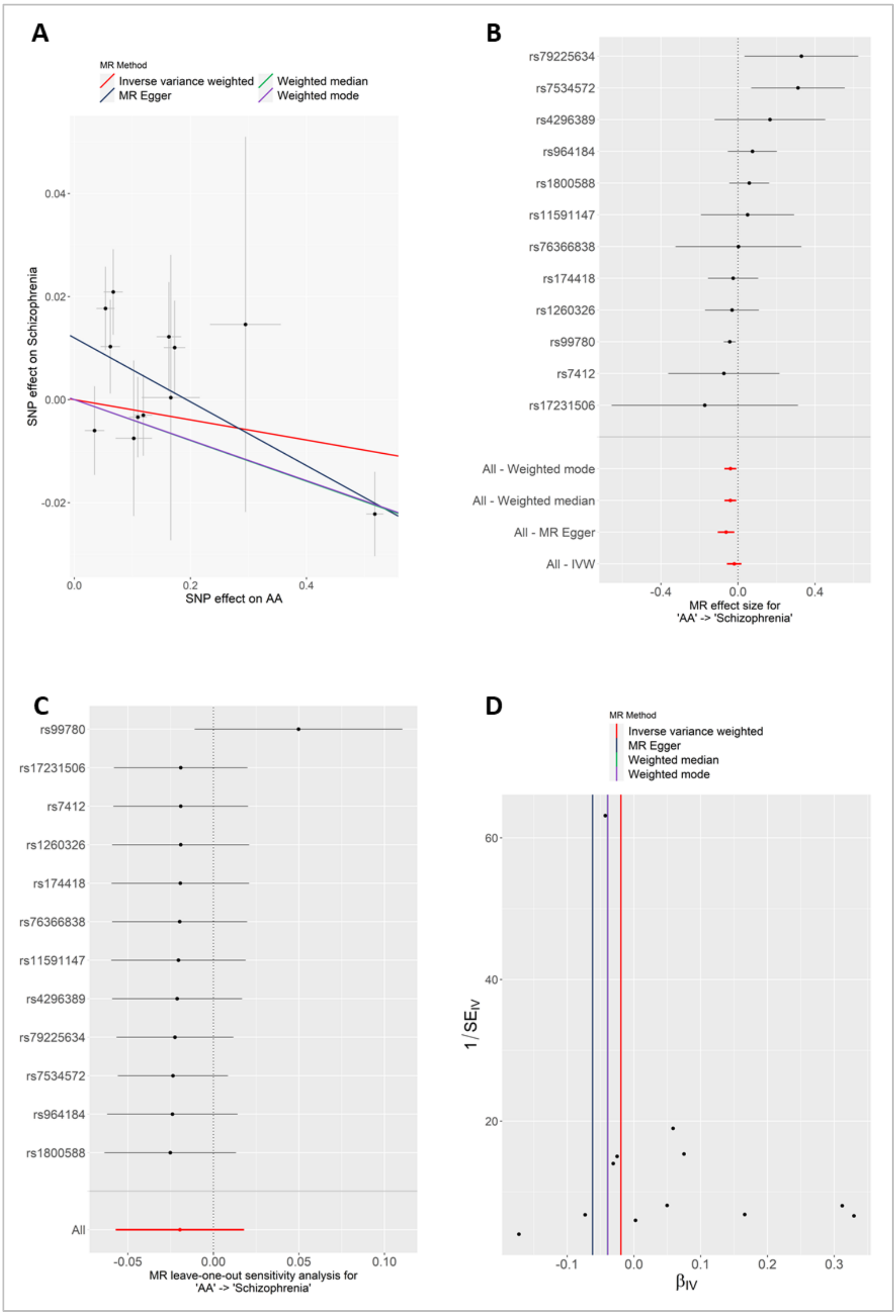


**Figure S6.** Mendelian randomization (MR) sensitivity plots of the causal effect of arachidonic acid (AA) levels on schizophrenia. A) Scatter plot showing results of the 4 MR multi-locus methods (see legend), B) Forest plot showing individual single nucleotide polymorphism (SNP) ratio estimates (SNP-outcome effect estimate / SNP-exposure effect estimate), C) Leave one out plot showing inverse variance weighted (IVW) estimates after omitting each SNP, and D) Funnel plot showing instrument precision: ratio estimate in log(odds ratio) ($\beta_{IV}$; x-axis) by instrument strength (1/ $\beta_{IV}$ standard error [SE]). Asymmetry in this plot may be a result of directional pleiotropy.

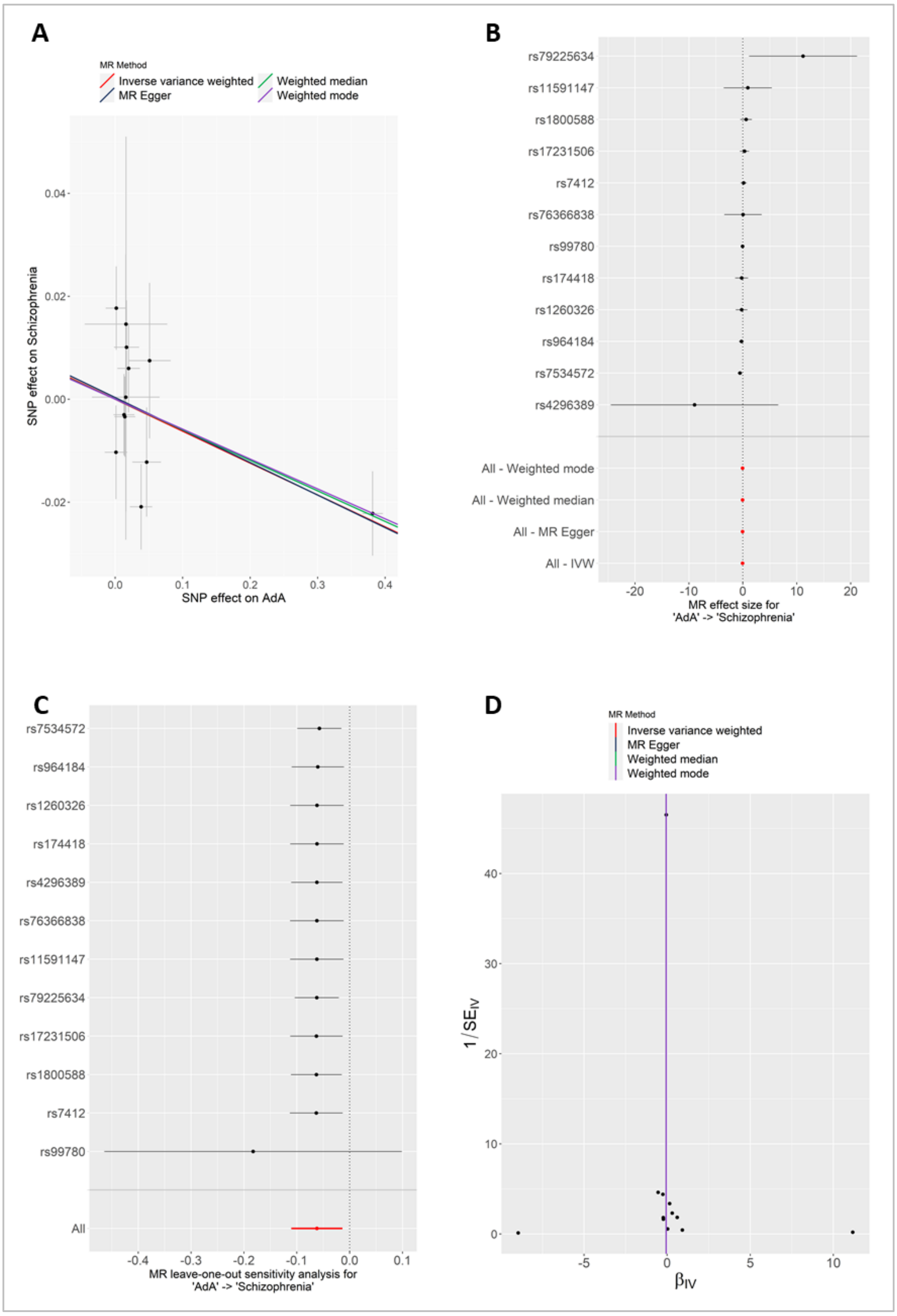


**Figure S7.** Mendelian randomization (MR) sensitivity plots of the causal effect of adrenic acid (AdA) levels on schizophrenia. A) Scatter plot showing results of the 4 MR multi-locus methods (see legend), B) Forest plot showing individual single nucleotide polymorphism (SNP) ratio estimates (SNP-outcome effect estimate / SNP-exposure effect estimate), C) Leave one out plot showing inverse variance weighted (IVW) estimates after omitting each SNP, and D) Funnel plot showing instrument precision: ratio estimate in log(odds ratio) ($\beta_{IV}$; x-axis) by instrument strength (1/ $\beta_{IV}$ standard error [SE]). Asymmetry in this plot may be a result of directional pleiotropy.
